# Process Model of Emotion Regulation-Based Digital Intervention for Emotional Problems — A mixed methods feasibility study

**DOI:** 10.1101/2022.11.24.22282695

**Authors:** Diyang Qu, Dongyu Liu, Chengxi Cai, Jiaao Yu, Quan Zhang, Kunxu Liu, Xuan Zhang, Ziqian Wei, Jiajia Tan, Zaixu Cui, Xiaoqian Zhang, Runsen Chen

## Abstract

The current study explored the feasibility of a newly developed self-guided digital intervention program TEA (Training for Emotional Adaptation) in alleviating depressive and anxiety symptoms, corresponding to the urgent call for remote mental health service. It is one of a few studies which adapted from theoretical models with effective intervention techniques. The first part involved 11 professional mental health practitioners giving feedback on the feasibility; while the second part involved a single-arm study with 32 participants recruited online, who went through the seven intervention sessions. The questionnaires were collected before, after, 14-days after and 30-days after the intervention. Moreover, 10 participants were invited to semi-structured interviews. Practitioners thought the TEA showed high professionalism (8.91/10) and is suitable for treating emotional symptoms (8.09/10). The Generalized estimating equation (GEE) model showed that the TEA significantly reduced their psychological symptoms, while the effects of the intervention were retained for 30-day post intervention (Cohen’s d >1). Thematic analysis revealed three main themes about future improvement, including content improvement, interaction improvement and bug-fixing. Taken together, current study supported the effectiveness of TEA, with the potential to address the urgent need for remote mental health service. Future randomized controlled trials to confirm the effectiveness are required.

## Introduction

Psychological distress refers to an individual’s negative emotional reaction to the stressors, including depressive and anxiety symptoms (Price & Woody, 2022). It has become a serious public health problem and causes an enormous burden for youth and the society in recent years. For example, the prevalence of depressive and anxiety symptoms increased sharply during the duration of COVID-19 pandemic (CMDC, 2021; WHO, 2022). In China, the prevalence of mental health problems has reached 17.5% (Xiang et al., 2022). However, the availability of mental health service is far from sufficient for individuals with emotional problems especially in developing countries, in which 79-93% of individuals with depressive symptoms, and 85-95% individuals with anxiety symptoms are having difficulties accessing effective mental health intervention (Chisholm et al., 2016). Therefore, it is of great importance to develop mental health interventions that are more cost-efficient and easier to access.

The increasing penetration of internet and smartphone provides an opportunity to increase the availability of mental health services through smartphone-based online platforms (Andrews et al., 2010). Especially during COVID-19 pandemic, the feasibility of in-person mental health services has been repeatedly reduced, while online digital mental health intervention (DMHI) provides us with an alternative (Liu et al., 2020).

Previous studies have validated the effectiveness of online DMHI for emotional problems and suggested that techniques from cognitive behavioural therapy (CBT) and mindfulness based cognitive behavioural therapy (MBCT) are effective in treating emotional disorders digitally (Fu et al., 2020; Gan et al., 2021). Among the existing DMHI for emotional disorders, the frequently involved intervention techniques include psychoeducation, behavioral activation, relaxation techniques, cognitive restructuring, mindfulness (Arjadi et al., 2018; Burton et al., 2016; Knaevelsrud et al., 2015; Moeini et al., 2019; Tiburcio et al., 2018; Yang et al., 2019).

However, the majority of the existing DMHI for emotional disorders in developing countries are mainly developed by simply combining several techniques (Arjadi et al., 2018; Burton et al., 2016; Tulbure et al., 2015; Yang et al., 2019; Wang et al., 2013), which might not be able to address emotional disorders from a more comprehensive perspective. Moreover, this simple combination of intervention techniques might result in less coherently structured session plans, which reduces the acceptability of the intervention (Sekhon et al., 2017). Therefore, it is necessary to develop a theory-based DMHI intervention that target for the emotional problems.

Moreover, some of the existing DMHI programs still involve mental health practitioners to offer additional support either online or in-person (Tiburcio et al., 2018; Arjadi et al., 2018; Marasinghe et al., 2012). Although the involvement of mental health practitioners can ensure the effectiveness of the intervention, the high-cost and high requirement of stand-by mental health practitioners are still not suitable for developing countries, due to the lack of enough mental health practitioners. Therefore, a cost-effective self-guided intervention program might be more suitable for meeting people’s mental health needs in developing countries.

### The Training for Emotional Adaptation (TEA)

To address the issues in the existing DMHI programs, we developed an emotion regulation model-based digital self-guided intervention for emotional disorders. The Training for Emotional Adaptation (TEA) is a self-help DMHI program, which merged effective CBT techniques with the process model of emotion regulation. The process model of emotion regulation describes emotion regulation is consisted of five sequential stages, including 1) situation selection, in which an individual actively selects situations that does not generate undesirable emotions; 2) situation modification, in which an individual alters the current situation to modify its emotional impacts; 3) attentional deployment, in which an individual directs one’s attention to stimuli that does not generate undesirable emotions; 4) cognitive change, which involves cognitive reappraisal of the already received stimuli to make it less disturbing; and (5) response modulation, in which an individual applies specific actions to control the already-triggered emotions (Gross, 2014). The process model of emotion regulation is of great importance in elucidating the whole process of emotion regulation and has been identified as effective in guiding intervention development (Bettis et al., 2022; Sakiris & Berle, 2019; Webb et al., 2012).

The TEA is consisted of seven sessions instructed online through WeChat mini program, in which the structure of the seven sessions follow the whole process model of emotion regulation. Following the themes of each stage of the process model, a series of CBT and MBCT techniques were assigned to each session, including psychoeducation, mindfulness, behavioral activation, worry tree, grounding techniques, self-compassion, savoring, cognitive restructuring and STOPP technique. The intervention was delivered online through the combination of verbal and graphical information, intervention videos, audio instructions and interactive practices.

### Aims

The current study aimed to investigate the feasibility and preliminary effect of the TEA program for emotional problems including depression and anxiety symptoms. We hypothesized that participants would improve on measures of symptoms of depression and anxiety. We also explored effects on sleep quality and emotion regulation strategies as well as the participants’ satisfaction and subjective evaluation of the intervention.

## Methods

### Sample

The current study was consisted of two parts. The first part involved collecting professional mental health practitioners’ feedbacks regarding the feasibility of the TEA as an online intervention for emotional disorders. The second part involved a single arm study to test the effectiveness of the TEA on alleviating emotional disorder symptoms.

For the professional’s feedback, our final sample involved 11 professional mental health practitioners as participants. These participants first filled an online questionnaire regarding to their demographic information. A brief introduction of the TEA along with potential feedback questions were then presented to the participants. Then participants went through the contents of the TEA WeChat mini program in two days, followed by an online questionnaire measuring participants’ opinions about the TEA.

For the single arm study, our final sample involved 32 participants, aged 18-36 (mean age = 25.38). Prior power analysis was conducted through G-power, which showed that to reach a statistical power of 85% with an effect size of 0.3, the sample size should be 31 or more. The eligibility criteria include 1) Participants aged 18-39; 2) Participants who can participate in our study through smartphones or other electronic devices; 3) Participants who are identified as having mild or more severe depression or anxiety symptoms by Patient Health Questionnaire (PHQ-9) or Generalised Anxiety Disorder (GAD-7) questionnaire (PHQ-9 >5 or GAD-7 >5). The Exclusion criteria includes 1) Participants who refused to give consent; 2) Participants who report having suicide risk within 1 week prior to the start of our study; 4) Participants who report having been diagnosed with psychotic disorders (e.g., schizophrenia, schizophrenic disorders), bipolar disorders, or other disorders co-occur with schizophrenic disorders; 5) Participants who report receiving other psychological intervention programs. Please refer to Figure 1 for the flow of participant recruitment.

**Figure 1.**
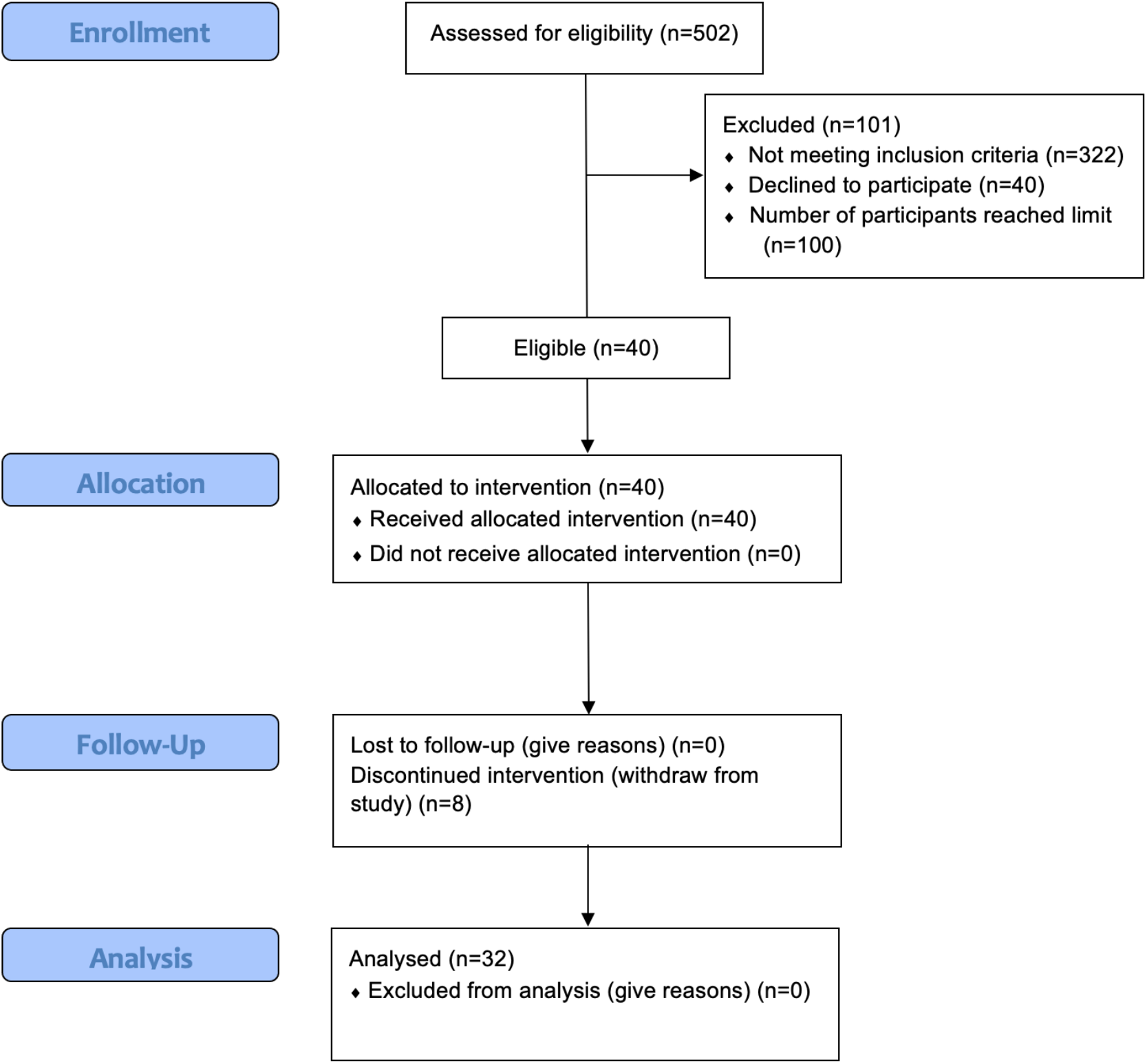
The Flow Chart of Participant Recruitment.

### Data collection

Participants were recruited in July 2022 via the flyer distributed through online poster. They indicated their interests in participation through scanning the Quick Response code provided with the flyer and completed the online eligibility test questionnaire. Participants who met the eligibility criteria were contacted by researchers. A short video of user’s guide along with a brief instruction of the study procedure were provided before participation. Then participants completed the pre-test questionnaire online and started participation in the TEA. The TEA involves seven sequential sessions, in which the participants were required to complete the seven sessions in 14 days, while participants should complete no more than one session in one day. Upon the completion of the seventh session, participants completed the post-test questionnaire online, and indicated their interests in participating in a voluntary one to one semi-structured interview. On the 14^th^ day after completion of the seventh session, participants completed the 14-day follow-up questionnaire online, while 10 participants were interviewed by team members online with experience with qualitative methods (i.e., semi-structured) through video conferencing software (TencentMeeting Video Communications) regarding to “*your experiences and feelings about the TEA for future improvements*”. Investigators explained the study’s aim, and all the content were recorded by audio (i.e., after obtained the consent from participants) then transcribed verbatim by a transcription service provided by Tencent Meeting and deidentified by interviewers later.

On the 30^th^ day after completion of the seventh session, participants completed 30-day follow-up questionnaire online. In total, 50 CNY (1 CNY = 0.14 USD) was given to each participant who completed the whole research process, to increase their intention of participation (See Figure 2 for the flow of this study).

**Figure 2.**
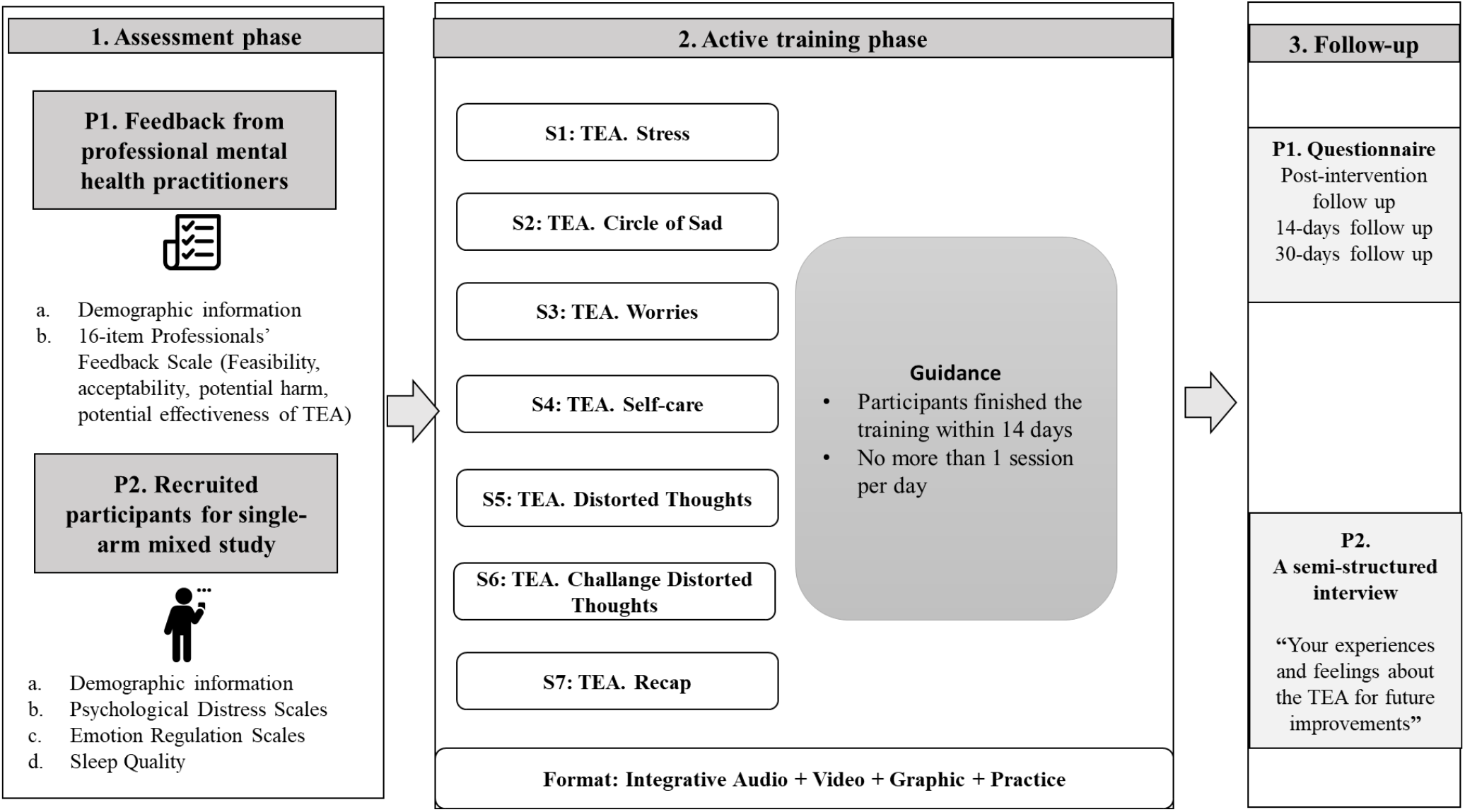
The Flow Chart the Current Study.

### Ethical Considerations

The current study received ethical approval from [BLINDED FOR REVIEW]. All participants were debriefed about the study and gave consent online before participation, while they were aware of their rights to withdraw at any point of the study with no harm. For participants who indicated recent suicidal ideation or suicide attempt, we provided professional help-seeking information when removing them from participation. All data was safely stored in a password-protected hardware, with only the relevant researchers have access to it. All personal information was kept confidential. Participants were given unique ID numbers for participant identification throughout the study.

### Measurements

#### Demographic Information

For the professionals’ feedback, participants self-reported their demographic information prior to the introduction to the TEA through online questionnaires. That includes age, gender, education level, professional title, and mental health-related work experiences. For the single arm study, participants self-reported their demographic information at eligibility test and pre-test. That includes age, assigned gender, gender minority, education level, socio-economic status, occupational status, marital status, religion, ethnicity, psychiatric diagnosis, current situation regarding to COVID-19-related restrictions and previous experiences of DMHI.

#### Professionals’ Feedback

Professional mental health practitioners’ opinions of the TEA were measured through a 16-item Professionals’ Feedback Scale specifically designed for the current study. That includes participants’ opinions about the feasibility, acceptability, potential harm, and potential effectiveness of the TEA. Participants self-reported their opinions through a 11-point Likert scale for each item ranged from 0 (e.g., “Very ineffective”) to 10 (e.g., “Very effective”).

#### Psychological Distress

Depressive symptoms were measured through the 9-item Patient Health Questionnaire (PHQ-9; Kroenke et al., 2001) at eligibility test, post-test, 14-day follow-up test and 30-day follow-up test. Participants self-reported their depressive symptoms through a 4-point Likert scale for each item ranged from 0 (“Not at all”) to 3 (“Nearly every day”), with a maximum possible total score of 27. The recommended cut-off point for clinically significant depression was set at 9 (Rizzo et al., 2000), while a score below 5 was indicated as not having depression symptoms (Kroenke et al., 2001). The PHQ-9 has good reliability and validity for measuring depressive symptoms in Chinese people (Wang et al., 2014).

Anxiety symptoms were measured through the 7-item Generalised Anxiety Disorder (GAD-7; Spitzer et al., 2006) questionnaire at eligibility test, post-test, 14-day follow-up test and 30-day follow-up test. Participants self-reported their anxiety symptoms through a 4-point Likert scale for each item ranged from 0 (“Not at all”) to 3 (“Nearly every day”), with a maximum possible total score of 21. The recommended cut-off point for clinically significant depression was set at 8 (Plummer et al., 2016), while a score below 5 was indicated as not having anxiety symptoms (Spitzer et al., 2006). The GAD-7 has good reliability and validity for measuring anxiety symptoms in Chinese people (Zeng et al., 2013).

#### Emotion Regulation

Emotion regulation abilities were measured through the 6-item short Emotion Regulation Questionnaire (ERQ-6; Gullone & Taffe, 2012) at pre-test, post-test, 14-day follow-up test and 30-day follow-up test. The ERQ-6 was developed through extracting 3 items from the cognitive reappraisal domain and 3 item from the expression suppression domain of the original Emotion Regulation Questionnaire (Gross & John, 2003). Participants self-reported their emotion regulation abilities through a 5-point Likert scale for each item ranged from 1 (“Strongly disagree”) to 5 (“Strongly agree”), with a maximum possible total score of 15 for each domain. The ERQ-6 has been shown as having good reliability and validity for measuring emotion regulation abilities (Gullone & Taffe, 2012).

#### Sleep Quality

Insomnia was used as a reversed indicator of sleep quality. Insomnia was measured through the 7-item Insomnia Severity Index (ISI-7; Bastien et al., 2001) at pre-test, post-test, 14-day follow-up test and 30-day follow-up test. Participants self-reported their insomnia severity through a 5-point Likert scale for each item ranged from 0 (e.g., “No problem”) to 4 (e.g., “Very severe problem”), with a maximum possible total score of 28. The cutoff point for having insomnia symptoms was set as 8 (Bastien et al., 2001). The ISI-7 has been shown as having good reliability and validity for measuring sleep quality in Chinese people (Yu, 2010).

### Statistics

All quantitative statistics were conducted using R-4.1.0 (R Core Team, 2017). Descriptive statistics were conducted for professionals’ feedback and demographic information for participants in the focus group interview and the single arm study separately. Descriptive statistics were also conducted for each item of the program feedback scale and the professional feedback scale. We calculated mean, standard deviation, median and range for continuous variables, and percentages for categorical variables.

We applied generalised equation (GEE) models to explore the predictive effects of measurement time-points and depression symptoms, anxiety symptoms, cognitive reappraisal emotion regulation, expression suppression emotional regulation and sleep quality, which reflect the impacts of the TEA intervention on these mental health characteristics.

Demographic information was controlled for in the GEE models. Post-hoc within-participant t-tests were conducted to explore the differences between theses mental health status at four time-points, which provides us with additional information about when the intervention impacts emerged and how long they remained. Bonferroni corrections were applied to reduce the risk of false positive.

For qualitative statistics, detailed verbatim transcriptions were refined and cross-checked by three researchers (CX; JA; LDY). Word software was used for coding and analysis of transcripts. The thematic analysis method was followed the steps proposed by Clarke et al. (2015), and following by one question, “*how to future improve the TEA from your own experience*”. Taken together, (1) For team members, including 2 who had previous experience with qualitative methods (CX; JA), familiarization with the data, independently reviewed the two transcripts and created an initial codebook (framework), by using comparison and consensus. (2) Then, the codebooks were collaborated and refined by four team members (CX; JA; LDY; ZX), any disagreement and possible solutions were identified and evaluated at research group meeting. Codes were grouped into themes iteratively until without further modification. (3) Then, themes about the *“future improvements of the TEA”* were extracted and refined by two researchers from all transcripts (CX; JA), and cross-checked by two researchers (LDY; ZX); (4) Examples were translated and double checked by researchers who are fluent in both Mandarin and English (CX; LDY).

## Results

Descriptive statistics can be found in table 1. Our final sample includes 32 participants in the single-arm study (mean age = 25.38, 87.5% female, 6.3% gender minority, 40% students, 6.3% with previous psychiatric diagnosis), and 11 participants in the focus group interview (mean age = 37.73, 63.6% female, mean year working in mental health service =12.55).

**Table 1.** Demographic Information of the Participants.

|  | TEA Participants (N=32) | Professional Mental Health Practitioners (N=11) |
| --- | --- | --- |
| **Age** |  |  |
| Mean (SD) | 25.38 (5.34) | 37.73 (6.68) |
| Median [Min, Max] | 23 [18, 36] | 39 [28, 51] |
| **Assigned Gender** |  |  |
| Female | 28 (87.5%) | 7 (63.6%) |
| Male | 4 (12.5%) | 4 (36.4%) |
| **Gender Minority** |  |  |
| No | 30 (93.8%) | - |
| Yes | 2 (6.3%) | - |
| **Educational level** |  |  |
| Mean (SD) | 6.63 (1.43) | 8.00 (1.55) |
| Median [Min, Max] | 6 [5, 10] | 8 [6, 10] |
| **Occupational Status** |  |  |
| Full-time Student | 13 (40.6%) | - |
| Full-time Employed | 13 (40.6%) | - |
| Part-time Employed | 2 (6.3%) | - |
| Unemployed | 4 (12.5%) | - |
| **Marital Status** |  |  |
| Unmarried | 27 (84.4%) | - |
| Married | 4 (12.5%) | - |
| Divorced | 1 (3.1%) | - |
| **Religion** |  |  |
| Buddhism | 4 (12.5%) | - |
| Christian | 1 (3.1%) | - |
| Atheist | 27 (84.4%) | - |
| **Ethnicity** |  |  |
| Han | 32 (100%) | - |
| **Socio-Economic Status** |  |  |
| Mean (SD) | 4.09 (1.86) | - |
| Median [Min, Max] | 4 [1, 6] | - |
| **Previous Psychiatric Diagnosis** |  |  |
| Yes | 2 (6.3%) | - |
| No | 30 (93.7%) | - |
| **Under COVID-19 Quarantine During the Study** |  |  |
| Yes | 1 (3.1%) | - |
| No | 31 (96.9%) | - |

**Table 1. Demographic Information of the Participants**
|  | TEA Participants (N=32) | Professional Mental Health Practitioners (N=11) |
| --- | --- | --- |
| **Previous Experiences of DMHI** |  |  |
| No Previous Experience | 32 (100%) | - |
| **Years in Mental Health Service** |  |  |
| Mean (SD) | - | 12.55 (7.84) |
| Median [Min, Max] | - | 14 [0, 27] |
| **Professional Title** |  |  |
| Mean (SD) | - | 2.73 (0.90) |
| Median [Min, Max] | - | 3 [1, 4] |
| **Subordinate Department in Hospital** |  |  |
| Psychological Crisis Intervention Center | - | 3 (27.3%) |
| Psychology Department (Counselling) | - | 2 (18.2%) |
| Psychological Intervention Center | - | 3 (27.3%) |
| Psychiatry Department | - | 2 (18.2%) |
| Clinical Psychology Department | - | 1 (9.0%) |

The results of the Professionals’ Feedback Scale are presented in Table 2. Overall, professionals’ evaluation of the TEA was positive. The professionalism of the TEA (i.e., the accuracy of how CBT techniques and illustrations were used) received the highest score (mean=8.92, SD=0.94), while the extent to which behavioural changes can be induced by the TEA received the lowest score (mean=7.18, SD=0.87).

**Table 2.**
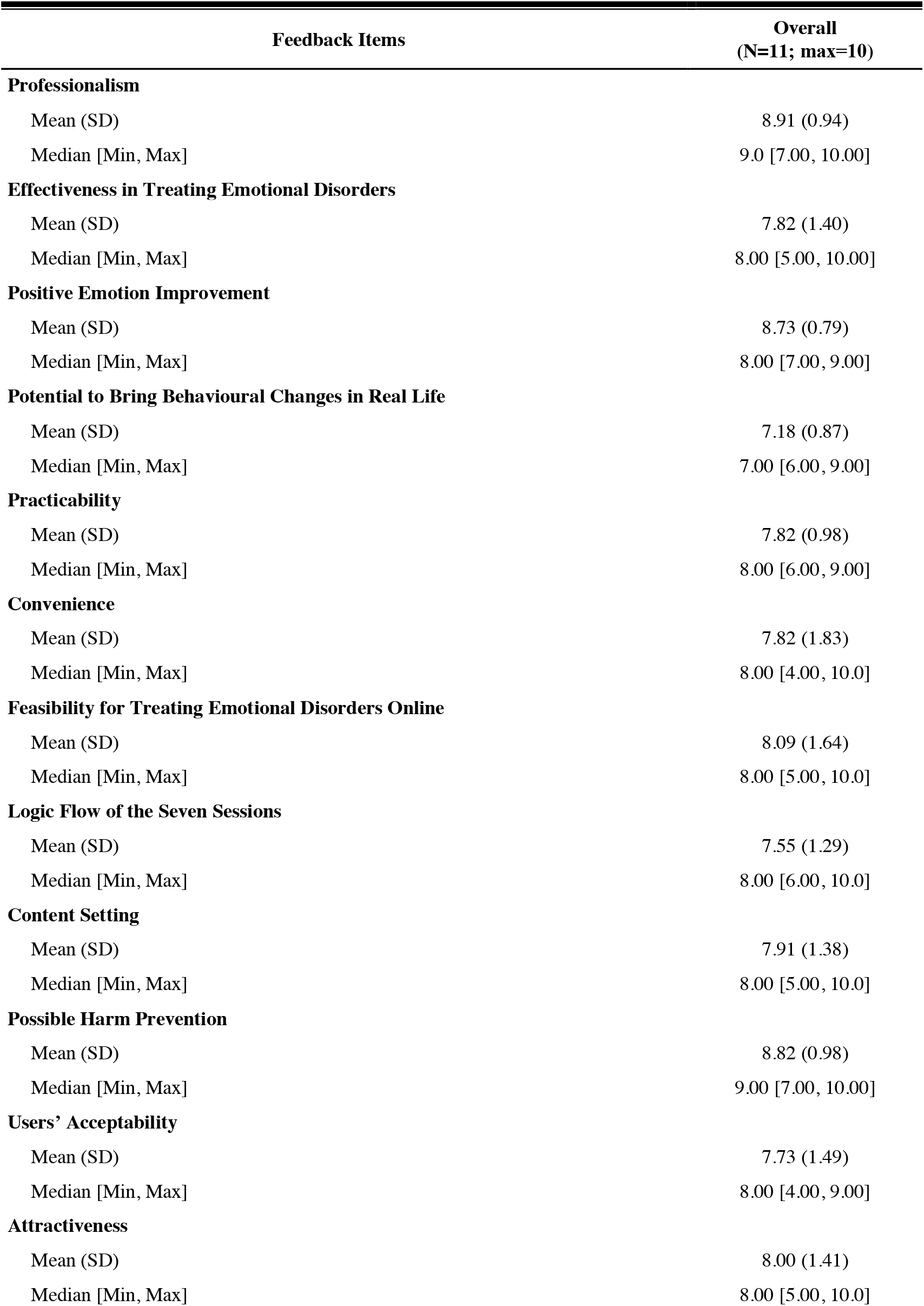
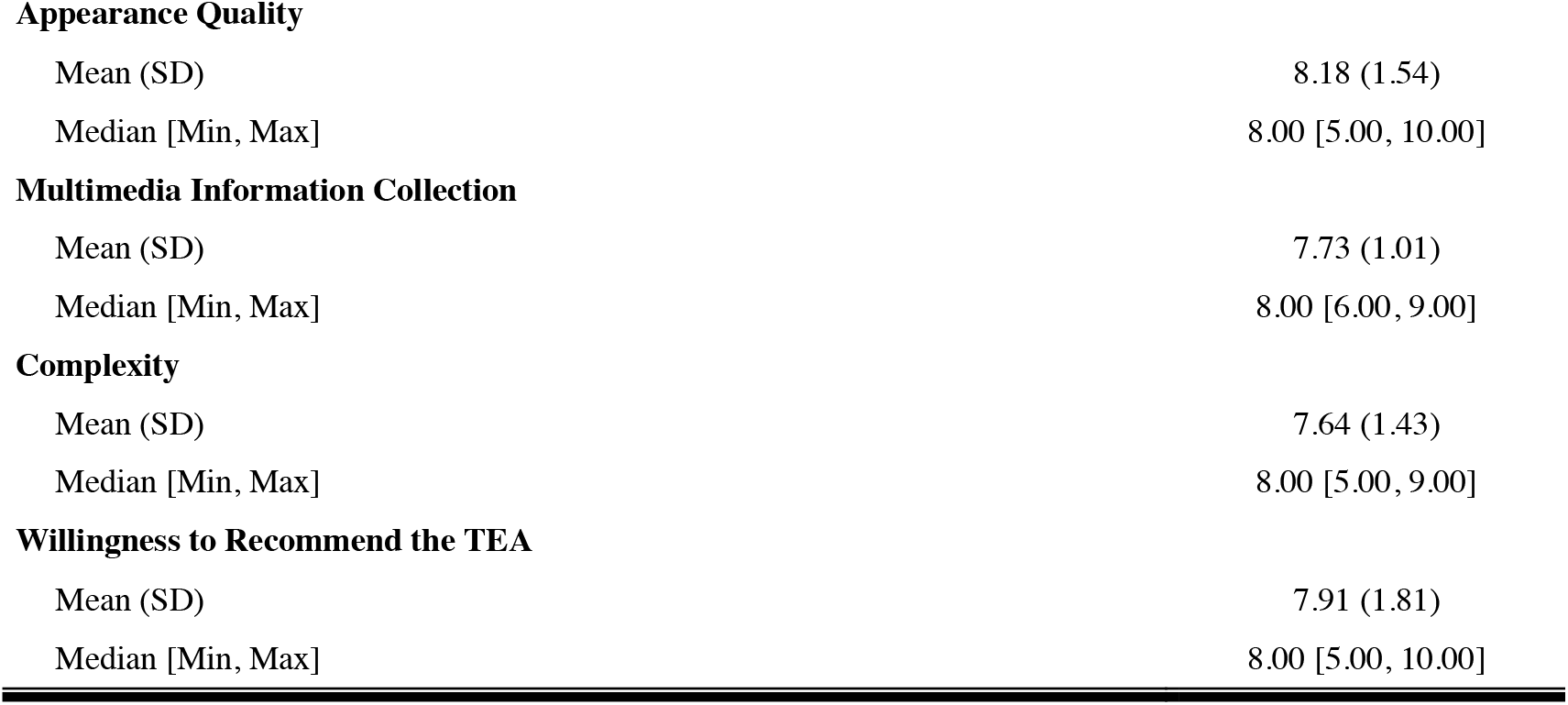
Training for Emotional Adaptation (TEA): Professional’s Feedback.

The results of the GEE model and post-hoc within-group t-tests are presented in Table 3. Generally, participants’ mental health symptoms reduced significantly after completing the TEA intervention, in which the symptoms levels remained low in a month after intervention (as illustrated in Figure 3).

**Table 3.** Intervention Effects of the Training for Emotional Adaptation (TEA)

| Outcomes | GEE |  | Post-hoc t-test (vs. pre-test) |  |  |
| --- | --- | --- | --- | --- | --- |
| | $\beta$ | SE | t-value (Cohen's D)<br>*post-test* | t-value (Cohen's D)<br>*14-day follow-up* | t-value (Cohen's D)<br>*30-day follow-up* |
| Depressive symptoms | -1.49*** | 0.30 | -4.29*** (0.76) | -4.85*** (0.86) | -6.37*** (1.13) |
| Anxiety symptoms | -1.46*** | 0.29 | -3.91** (0.69) | -4.55*** (0.81) | -6.45*** (1.14) |
| Cognitive Reappraisal | 1.63*** | 0.24 | -1.74 (0.31) | 4.67*** (0.83) | 5.03*** (0.89) |
| Expression Suppression | -2.03*** | 0.23 | 0.15 (0.03) | -7.61*** (1.34) | -6.12*** (1.08) |
| Insomnia | -1.32*** | 0.37 | -3.73* (0.66) | -4.02** (0.71) | -5.24*** (0.93) |

Notes. \*\*\*= $p<0.001$ ; \*\*= $p<0.01$ ; \*= $p<0.05$ ; GEE: Generalised Equation Modelling. Bonferroni Corrections were applied

**Figure 3.**
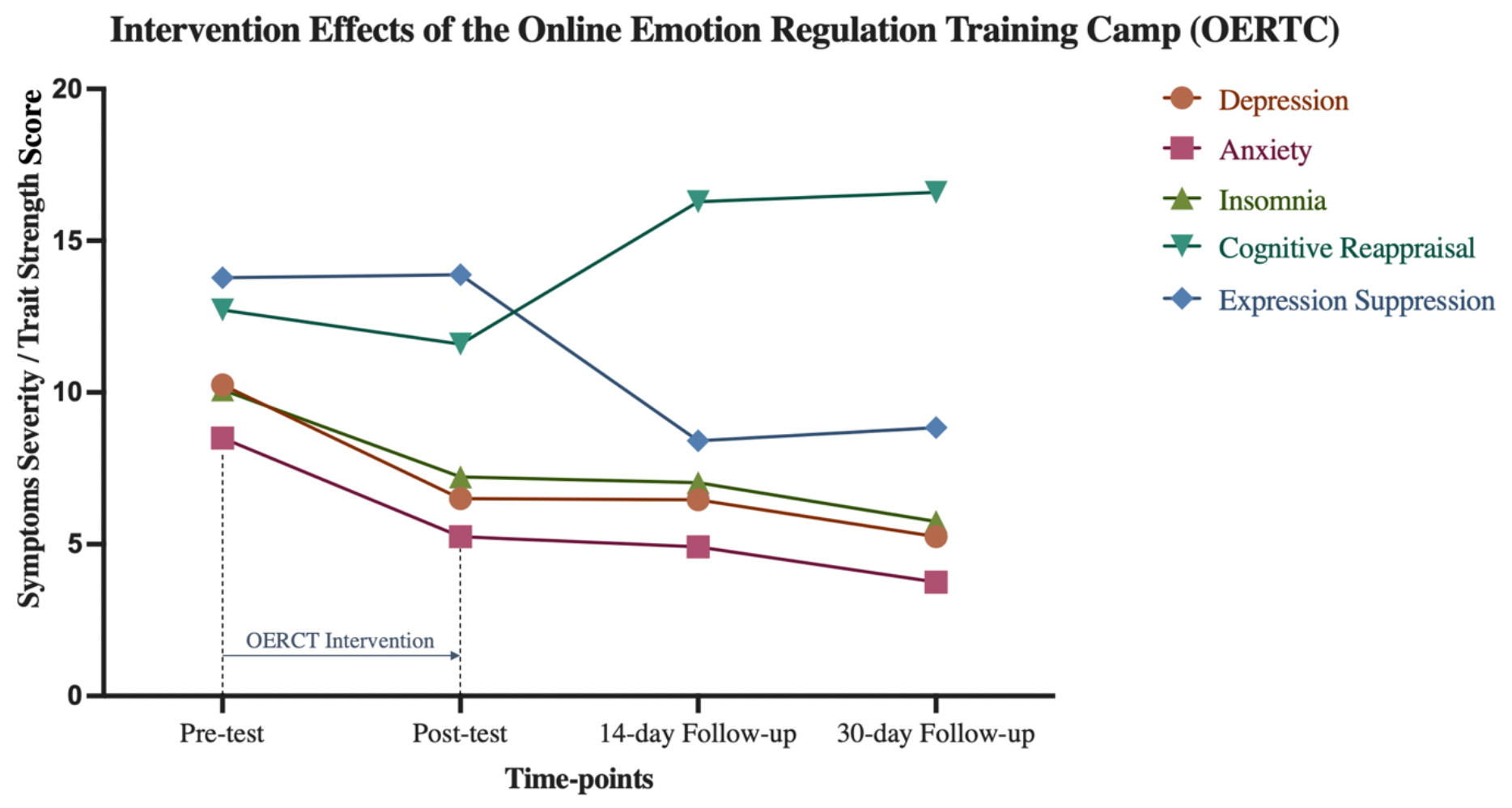
Intervention Effects of the Training for Emotional Adaptation (TEA)

Specifically, for depressive symptoms, the GEE model indicated significant intervention effects of the TEA on depression symptoms (β=-1.49, p<0.001). The post-hoc t-tests showed that compared to pre-intervention, depression symptoms significantly reduced after intervention (t=-4.29, p<0.001), while the intervention effect remained at 14-day follow-up (t=-4.85, p<0.001) and 30-day follow-up (t=-6.37, p<0.001).

For anxiety, the GEE model indicated significant intervention effects of the TEA on anxiety symptoms (β=-1.46, p<0.001). The post-hoc t-tests showed that compared to pre-intervention, anxiety symptoms significantly reduced after intervention (t=-3.91, p=0.007), while the intervention effect remained at 14-day follow-up (t=-4.55, p<0.001) and 30-day follow-up (t=-6.45, p<0.001).

For cognitive reappraisal emotion regulation, the GEE model indicated significant intervention effects of the TEA on cognitive reappraisal (β=1.63, p<0.001). The post-hoc t-tests showed that compared to pre-intervention, cognitive reappraisal did not significantly change after intervention (t=-4.29, p=1.365). However, cognitive reappraisal significantly increased at 14-day follow-up (t=4.67, p<0.001), while the intervention effect remained at 30-day follow-up (t=5.03, p<0.001).

For expression suppression emotion regulation, the GEE model indicated significant intervention effects of the TEA on expression suppression (β=-2.03, p<0.001). The post-hoc t-tests showed that compared to pre-intervention, expression suppression did not significantly change after intervention (t=0.15, p=13.23). However, expression suppression significantly decreased at 14-day follow-up (t=-7.61, p<0.001), while the intervention effect remained at 30-day follow-up (t=-6.21, p<0.001).

For sleep quality, the GEE model indicated significant intervention effects of the TEA on sleep quality (β=-1.32, p<0.001). The post-hoc t-tests showed that compared to pre-intervention, insomnia severity significantly reduced after intervention (t=-3.73, p=0.012), while the intervention effect remained at 14-day follow-up (t=-4.02, p=0.005) and 30-day follow-up (t=-5.24, p<0.001).

The results of thematic analysis of the semi-structured interview are presented in Table 4. Participants suggestions about future improvements of the TEA were classified into three themes, including “content improvement”, “interaction improvement”, and “bug-fixing”. Overall, participants’ suggestions are mainly about user experience. In the content improvement theme, participants suggested to improve user engagement through gamification and more encouraging reinforcement. Additional contents were suggested to include more detailed examples, review sessions and the marking completion section. Few participants also indicated desire of optional more detailed and difficult contents for people who find the existing contents too shallow. In the Interaction improvement theme, participants suggested to improve multimedia interactions and add daily reminders for completing the sessions. Participants also indicated desires of better communication experiences both with peers and with staff. In the Bug-fixing theme, participants mainly reported program crash and content saving problems that need to be fixed.

**Table 4.** Training for Emotional Adaptation (TEA): Participants’ Improvement Suggestions.

| Themes | Sub-themes | Codes | Significant Statement Examples<br>(Translated from Chinese) |
| --- | --- | --- | --- |
| Content Improvement | User Engagement | Gamification | "You can modify the existing contents into interactive games, like *super Mario* that involves different sessions and tasks." |
|  |  | Reinforcement / Reward | "The current virtual rewards given (upon completing each session) are not encouraging enough." |
|  | Additional Contents | Detailed Examples | "You can provide me with a detailed list that includes the activities I can do in a week..... so I know what (activities) to do to improve my emotional states." |
|  |  | Recap Sessions | "Although I have the willingness to change, I probably need to repeat (the training), or have recap sessions, as we only have seven sessions." |
|  |  | Marking Completion | "You can create a marking section for me to mark the activities I should complete, to help me make behavioural changes" |
|  | Individualization | Optional More Detailed and Difficult Contents | "If anything, I would like to have some deeper contents. I think the current content is relatively shallow. You can have the contents graded (from easy to difficult)." |
| Interaction Improvement | Operational Design | Accessibility | "It is sometimes not fluent when I want to flip the page. It would be better to just scroll all of the pages down." |
|  |  | Prevention User Errors | "It easy to accidentally tap on the slider button which will open a figure" |
|  | Multimedia-interaction | Audio-Verbal Information Matching | "I find the audio recording a bit distracting." |
|  |  |  | "The audio recording at one page sometimes contains information on the next page." |
|  | Notification | Daily Reminders | "I think it can push a notification, just to remind you to complete the session today." |
|  | Communications | Message Board | "To set up a message board, for us to record some of our puzzles, and then maybe we can answer them next time." |
|  |  |  | "To have interactions..... you can have your problems answered." |
| Bug-fixing | Program Crash | Program Crash when Flipping the Pages | “Every time I want to go back to the previous page, I slide back to the previous page from left to right, sometimes I find that I just quitted.” |
|  | Display Error | Input Box Error | “Sometimes the input box will jump to the top of the page.” |
|  | Saving of Completed Contents | Completed Contents Not Saved When re-logging in | “When I accidentally quitted the program, the contents I completed before will all disappear.” |

## Discussion

The current study verified the effects of the TEA on reducing emotional problems symptoms through combining quantitative data from a one-month single arm study and qualitative data from a professionals’ focus group and semi-structured interviews. Our results supported the feasibility and preliminary effectiveness of the TEA in treating emotional symptoms including depression and anxiety.

Our results from the single-arm study indicated a significant reduction of both depression and anxiety symptoms upon the completion of the intervention, in which the intervention effect lasted for a month. Our results supported the effectiveness of unguided digital intervention on emotional disorder symptoms in China, which is also in line with previous studies in other countries (Fairburn & Patel, 2017). Compared to other digital interventions for depression and anxiety, which generally showed small to moderate intervention effects (Harrer et al., 2019), The TEA showed moderate intervention effects for both depression (*d* = 0.76) and anxiety (*d* = 0.69) post-intervention. Moreover, comparing to pre-intervention, The TEA showed large intervention effects for both depression and anxiety at follow-up, which indicated a great potential for TEA to be applied as a self-guided digital intervention for depression and anxiety. From professionals’ feedback, the TEA also received high ratings for professionalism, practicability, effectiveness in treating emotional disorders, and feasibility for online intervention, which guaranteed its’ potential in treating depression and anxiety as a self-guided digital intervention program.

One of the strengths of the TEA is that it systematically combined different intervention techniques based on the whole process of emotion regulation (Gross, 2014), while aiming at alleviating depression and anxiety symptoms through intervening with a comprehensive aspect of factors related to emotion regulation. Our results also supported the impacts of the TEA on different factors including physiology and cognition. Physiologically, our results showed significant improvement in sleep quality after the intervention, while the intervention effect remained stable in follow-up. An abundance of previous studies have found significant associations between sleep quality and both depression and anxiety (Fang et al., 2019; Nyer et al., 2013; Alvaro et al., 2013). Previous studies have supported the effectiveness of digital CBT on insomnia comorbid with depression (Sadler et al., 2018; Cheng et al., 2019), which align well with our results showing reduced insomnia symptoms along with emotional problems. Through the combination of grounding techniques including relaxation trainning and mindfulness that have been shown to reduce sleep disturbance (Rusch et al., 2019; Liu et al., 2020), the TEA showed the potential to reduce emotional problem symptoms through inducing sleep quality. However, the underlying mechanisms from sleep quality to emotional problem symptoms still need to be verified by further investigations.

From the cognitive perspective, our results indicated a significant improvement in participants’ ability in cognitive reappraisal, which supported the impacts of the TEA on improving user’s emotion regulation as conceptualized in the process model of emotion regulation (Gross, 2004). One meta-analysis concerning the effective components of intervention programs developed based on the process model of emotion regulation indicated that cognitive change has the largest intervention effects among all aspects of the emotion regulation process (Webb et al., 2012). The TEA has two whole sessions concerning cognitive restructuring, which guaranteed its potential in reducing emotional symptoms through inducing cognitive reappraisal ability. However, one interesting finding is that the expression suppression was reduced in our participants. This went against previous studies indicating expression suppression as another emotion regulation strategy by previous studies (Gross, 2001; Gross & John, 2003). Nevertheless, expression suppression has also been identified as a maladaptive coping strategy that damages one’s mental health (Chervonsky & Hunt, 2017; Cutuli et al., 2014), which supported the role of the TEA in reducing emotional symptoms. Moreover, it has been shown that compared with expression suppression, cognitive reappraisal serves as a more healthier emotion regulation strategy (Cutuli et al., 2014; Ferschmann et al., 2021), which supports the intervention effects of the TEA. Another interesting point is that cognitive reappraisal and expression suppression did not show significant change right after the intervention, but significant changes showed in 14-day and 30-day follow-ups. This indicated delayed impacts of the TEA on emotion regulation strategies, which is also in line with previous studies finding cognition as stable and difficult to change (Clapp, 1993). This also supported the importance of incorporating a comprehensive process of emotion regulation in an intervention program. However, future studies are in need to examine the effects of different intervention components of the TEA.

While the TEA has been shown as an effective intervention program for depression and anxiety, there are certain limitations that we need to admit. First of all, the current study utilized a single-arm design, which diminished the strength of our evidence supporting the intervention effects of the TEA. Moreover, the current study failed to explore the impacts of each intervention components on mental health outcome, which means that we cannot draw a conclusion on whether the moderate-to-large impacts of the TEA on mental health outcomes is a result of an incorporation of techniques targeting the whole emotion regulation process. Therefore, the future randomized controlled trails with more comprehensive measures are still in need to further verify the intervention effects of the TEA on depression and anxiety. Moreover, the semi-structured interviews also indicated some limitations regarding the current version of the TEA, which call for further refinements of the program in future versions of the program, including having additional and individualized contents, improving users’ experiences through refining software interaction styles, as well as bug fixing. Despite these, the merit of the TEA should be strongly addressed; the enormous size of WeChat user (i.e., over 1.2 billion monthly active users), making our self-guided TEA far more cost-effective and implementation which are easier to service more people; especially about 10% of WeChat users are international users (Content, 2021), in this case, the approach may further help individual who are not currently living in China (e.g., Immigrants), and not used to get active use of Facebook, WhatsApp or Twitter.

## Conclusion

To address the current needs for digital mental health intervention programs to account for the insufficient availability of mental health service in China, the current study investigated the feasibility of the Online Emotion Regulation Training Camp (TEA) as a self-guided digital intervention for emotional disorders. Through the combinations of feedbacks from mental health professionals, a one-month single-arm study and semi-structured interviews, our results supported the TEA as an effective intervention program for depression and anxiety, while the intervention effects remain stable across a month. However, future RCT studies are still in need to further verify the effects of the TEA.

## Data Availability

All data produced are available online at https://osf.io/hpg4s/

## Notes

### Competing Interest Statement

The authors have declared no competing interest.

### Clinical Trial

ChiCTR2200065944

### Funding Statement

This study was funded by Vanke School of Public Health, Tsinghua University

### Author Declarations

Ethics committee of Vanke School of Public Health, Tsinghua University gave ethical approval for this work.

